# Evaluation of Direct susceptibility testing method for Moxifloxacin against Mycobacterium tuberculosis using the BACTEC MGIT 960 system

**DOI:** 10.64898/2026.03.07.26346296

**Authors:** Subhasish Bhadra, Ujjwala Gaikwad, Kumar Vikram, Sachin Chandrakar, Abhijit Kumar Prasad

**Author notes:** Corresponding Author: Dr. Ujjwala Gaikwad Professor, Department of Microbiology, All India Institute of Medical Sciences Raipur, Tatibandh, G.E. Road, Raipur 492099, India, Email id- Contact Number. Email id- Contact Number-; Email id- Contact Number-; Email id- Contact Number-; Email id- Contact Number-; Email id- Contact Number.

## Abstract

**Background:** Moxifloxacin is a key component of current MDR-TB therapy regimens. The choice to include it in therapy at standard or higher doses is based on the lack or presence of resistance mutations conferring low-level or high-level resistance to moxifloxacin, as detected by the Line probe assay (LPA). Due to inherent phenotypic and genotypic discordance, such resistance must be reconfirmed phenotypically using liquid culture and drug susceptibility testing (LC-DST) at critical concentration and clinical breakpoint of the drug. This takes several weeks, delaying the therapeutic decision. The current study intends to shorten this time by performing phenotypic DST directly on sputum samples.

**Methods:** A cross-sectional study was conducted for 18 months from October 2023 to April 2025, in which smear positive sputum samples that were resistant to Rifampicin or Isoniazid or both were subjected to Direct Moxifloxacin DST, irrespective of patient characteristics. Results obtained by Direct DST were compared against Indirect LC-DST as the gold standard as well as with LPA to evaluate the diagnostic accuracy and time savings with direct DST.

**Results:** Direct DST exhibited high accuracy of 98.18%, high sensitivity (90.91%), high specificity (98.99%), excellent concordance (98.18%) and almost perfect agreement (kappa value - 0.901) when compared to Indirect DST. It saved an average of 10 ± 3.20 days over Indirect DST to obtain the valid results. Similar performance was also observed in comparison to LPA with good sensitivity (90.91%), specificity (98.99%) and accuracy (98.18%). Significant discordance was however noted in classification of resistance by both direct and indirect DST compared to LPA. Few error rates and minimal cost advantages were some of the disadvantages of Direct-DST.

**Conclusion:** Direct DST demonstrated excellent performance characteristics, making it a reliable and rapid alternative to the gold standard, saving significant time in guiding therapeutic decisions for effective patient management.

## 1. Introduction

According to WHO global report 2025, tuberculosis is the world’s leading cause of death from a single infectious agent and among the top 10 causes of death worldwide^1^. The emergence of drug resistant TB (DR-TB) has posed great challenge to the global TB control measures. Early detection of resistance is extremely important for effective treatment and control.

With effectiveness comparable to isoniazid^2,3^, moxifloxacin (MFX) now forms the backbone of DR-TB treatment^4^ and has recently been introduced by WHO in shorter 4-month regimen for DS-TB^5^. It also continues to be part future treatment options under evaluation^6^.

According to the current diagnostic and treatment algorithm for TB followed in India, all rifampicin resistant (RR)/ Multi-drug resistant (MDR)/Isoniazid resistant (Hr) TB isolates diagnosed by GeneXpert and First line-Line probe assay (FL-LPA) are subjected to Second line-Line probe assay (SL-LPA) in order to find out mutations conferring resistance to the anti-TB Fluroquinolones (moxifloxacin and levofloxacin)^7^. Moxifloxacin can be administered at standard dose of 400 mg/day or at a higher dose of 800 mg/day, the choice to include it in therapy at standard or higher doses is based on presence or absence of resistance mutations conferring low-level or high-level resistance to moxifloxacin, as detected by the LPA^8,9^.

LPA enables faster diagnosis by detecting mutations. However, the genotypic tests have limitations resulting in high degree of genotypic and phenotypic discordance for moxifloxacin^10,11^. WHO thus recommend reconfirmation of fluroquinolone resistant strain detected by LPA, with liquid culture DST (drug susceptibility testing)^12^. Especially, it is always recommended to subject the isolates with low level MFX resistance to phenotypic DST by Mycobacterial Growth Indicator Tube (MGIT) at critical concentration (CC) (0.25 μg/ml) as well as clinical breakpoint (1.0 μg/ml) to assess whether higher doses of MFX will work in the absence of resistance at Clinical Breakpoint (CB).

Currently this phenotypic DST for MFX is done by indirect DST method in which sample is first decontaminated and cultured in MGIT and the growth obtained is further subjected to DST. Indirect DST takes double time (time to grow the organism + time to test the organism for DST) to obtain the final results. This results in huge delays to obtain confirmatory results and thus increases risk of treatment failure^10,12,13^.

Considerable amount of time can be saved if phenotypic DST at both CC and CB for MFX can be done directly from the clinical samples in parallel to LPA testing. Previous studies have attempted such kind of direct phenotypic DST using MGIT for rifampicin and isoniazid with encouraging results^13,14,15^.

However limited studies exist to evaluate the reliability, accuracy, time savings and cost savings associated with direct phenotypic DST against MFX.

The present study aims at standardizing and evaluating the diagnostic accuracy of ‘Direct DST’ method for moxifloxacin at both CC and CB and perform a comparative analysis with currently used standard drug susceptibility methods.

## 2. Materials and Methods

### 2.1. Study Design and Participants

A prospective diagnostic accuracy study was conducted in the Department of Microbiology, All India Institute of Medical Sciences, Raipur, Chhattisgarh and Intermediate Reference Laboratory, Lalpur, Raipur, Chhattisgarh. In a cross-sectional design, the study was conducted for 1 year 6 months (October 2023-April 2025).

Study participants included patients of all age groups irrespective of treatment status whose sputum samples were received at the mycobacteriology laboratory as a part of standard treatment care, were diagnosed as tuberculosis on initial CBNAAT/Truenat testing and additionally smear positive irrespective of degree of smear positivity. Such patients were traced back from the laboratory to obtain the clinical details. A second specimen was collected from such patients and subjected to Direct FL-LPA, specimens resistant of either Isoniazid, Rifampicin or both were consecutively recruited. The study was reported in accordance with the STARD guidelines^16^.

### 2.2. Study Procedure

Study samples were subjected to LPA for SL drugs and Indirect DST to MFX at CC (0.25 μg/ml) and CB (1.0 μg/ml) as per standard laboratory protocol and Direct DST to MFX at CC (0.25 μg/ml) and CB (1.0 μg/ml) as per current study protocol, the results were compared with the standard methods. Direct DST method was standardized before processing the patient samples.

#### 2.2.1. Preparation of Drug solution

Drug powder of Moxifloxacin was obtained from the national reference laboratory under National Tuberculosis Elimination Programme (NTEP) network. Drug powder used was Moxifloxacin hydrochloride VETRANAL™ by SUPELCO, analytical standard. Drug solution was prepared using the formula-

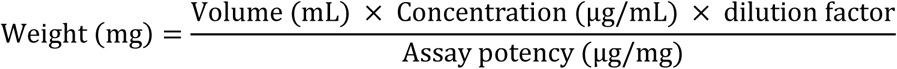

Stock solution was prepared 100 times the concentration of the working solution, it was aliquoted in 120 μL volume and stored at -20 °C for 1 year. The working solution was prepared by making 1:100 dilution of stock solution and stored at [2°C to 8 °C] for upto 1 week.

#### 2.2.2. Indirect DST Procedure

Decontaminated samples in 0.5 ml amount were inoculated in the fresh MGIT tube containing PANTA and supplement and incubated in BACTEC MGIT 960 system. The tubes once flagged positive were confirmed for growth of MTBC by performing ZN staining and MPT-64 antigen assay using (Bioline™ TB Ag MPT64 by Abbott). Inoculum from positive tubes without contamination were subjected to indirect DST against second line drugs including Moxifloxacin as per the standard protocol^12^- After positive growth signal (day 0), the MGIT growth tube was incubated upto day 5 before using the tube for indirect DST [tubes were diluted at 1:5 ratio at or after Day 3]. The growth in MGIT was now inoculated to drug containing MGIT tubes (at both CC and CB) and GC (Growth control) tube which was prepared by diluting the inoculum in 1:100 dilution. The tubes were added with ODAC supplement without antimicrobial mixture (PANTA) and incubated with a 4 to 13 days protocol.

#### 2.2.3. Direct DST procedure

A modified protocol compared to standard indirect DST was developed. Modifications included- (i) A extended 4-21 days incubation protocol was adopted. (ii) Dilution factor of 1:10 was used for growth control (iii) The antimicrobial mixture PANTA (Becton Dickinson Diagnostic Systems, Sparks, MD) was added to the control as well as the drug containing MGIT tubes to suppress contamination.

#### 2.2.4. Standardization of Direct DST

Since the method under evaluation in this study is a novel method for MFX susceptibility testing, the Direct DST protocol was standardized using the samples containing MTBC isolates with known susceptibility pattern. For this, 10 sputum samples that have been already subjected to direct LPA for first and second line drugs with known results were obtained from the reference laboratory. The investigators were blinded towards their susceptibility pattern. The samples were divided in two parts and testing was performed on each isolate in duplicates to assess the reproducibility.

#### 2.2.5. Quality Control of Direct DST

*M. tuberculosis* H37Rv strain (ATCC 27294) was used for quality control (QC) testing in DST. This strain was introduced at each time when a batch of DST was set up or as every 6th isolate in a run. If any resistance in the QC strain is observed, all the other results in that batch were considered invalid and subjected to repeat testing. However, Standard drug resistant strain of MTBC were not available for QC testing of resistance.

#### 2.2.6. Interpretation of DST

Test was considered valid if control tube reached growth unit (GU) of 400 units in 4-21 days. If it reached the value before 4 days it resulted in X400 error, if it did not reach 400 GU in 21 days it results in X200 error- No interpretation can be done in such scenarios. After control reaches 400 GU, the drug tubes were interpreted. If the drug tube reached a GU value of >100 GU then it was considered resistant at that concentration of drug in the tube, if it was <100 GU it was sensitive at that concentration.

#### 2.2.7. Line Probe Assay

The first-line LPA was carried out using MTBDRplus version 2.0 kit (Hain Life Sciences Pvt. Ltd). Samples with valid results that had resistant mutations were subjected to second-line LPA using MTBDRsl version 2.0 kit (Hain Life Sciences Pvt. Ltd). Processing and interpretation of LPA was done with extracted DNA according to the manufacturer’s instructions^9^.

#### 2.2.8. Data Analysis

The data was collected and entered in Microsoft excel on regular basis and analysed using SPSS version 21. Results obtained for moxifloxacin susceptibility by Direct DST was compared against Indirect LC- DST considering Indirect DST as the gold standard as well as findings on Line probe assay testing. Outcome variables including sensitivity, specificity, PPV, NPV, diagnostic accuracy, concordance and discordance of Direct DST against Indirect DST and LPA; time saving and cost effectiveness of direct DST were also analysed. The analysed data was presented in the form of frequency distribution/Proportions and Graphs for qualitative data, while quantitative data was presented in the form of Mean ± SD. The statistical significance was calculated by using Chi-square test or other appropriate statistical methods according to the type of data obtained. P value of <0.05 was taken as measure of statistical significance.

## 3. Results

A total of 110 samples showed valid interpretable results by both direct and indirect DST out of total 131 samples processed. Remaining 21 samples were excluded from the analysis since they showed X400 and X200 errors during incubation in MGIT (fig.1).

**Figure 1.**
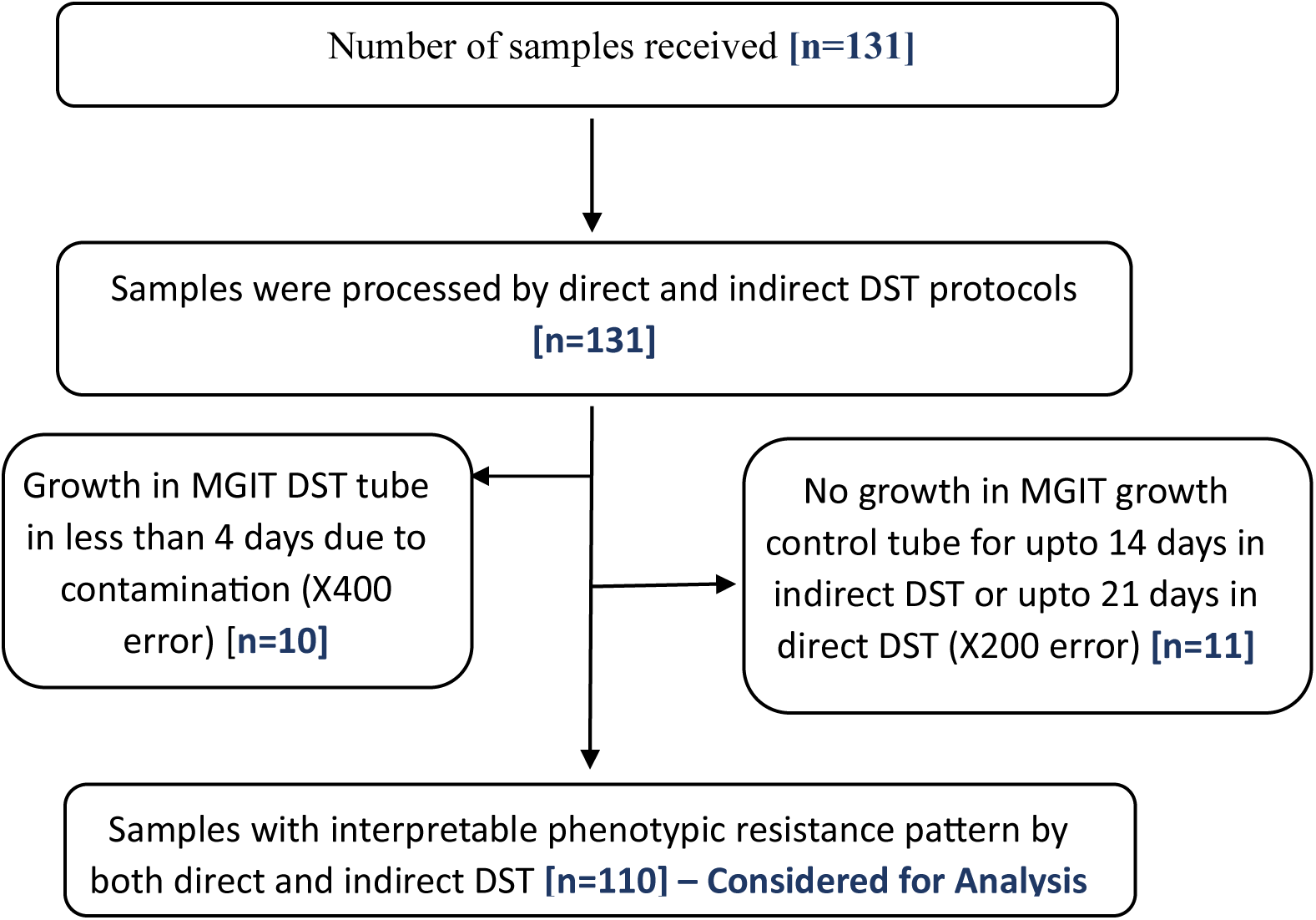
Patient Recruitment and workflow

### 3.1. Patient parameters

Majority of the patients were of young age group (15-47 years) (67.27%) and had male gender (69.09%).

Majority (n=87) patients were new cases, who never had treatment for TB or had taken anti-TB drugs for less than one month and while 23 patients were previously treated cases.

### 3.2. Drug resistance profile of the patients

All the patients had resistance to at least one first line anti-TB drug as per the inclusion criteria. However, majority (N=100) showed resistance to isoniazid which comprised of isoniazid monoresistance in 80.91% cases and total isoniazid resistance (Monoresistant + MDR) of 90.9%, among which most of them had high level resistance to isoniazid(N=71). Rifampicin monoresistance was noted in 9.09 % cases (N=10) and total rifampicin resistance (Monoresistance + MDR) was noted in 19.09 % cases (N=21). While, 10% samples were MDR that were resistant to both isoniazid and rifampicin (N=11). Resistance rate for Fluroquinolones among the first line resistant samples was 10% (11 out of 110) by LPA which included five samples with high level resistance to Moxifloxacin, five samples with low level resistance mutation to Moxifloxacin. One sample had unknown mutation and the resistance was inferred which resulted in labelling it as having at least low level resistance. When FQ resistance was studied in reference to resistance to first line drugs, it was noted that a significant proportion of MDR isolates were resistant to fluroquinolones-45.45% (5 out of 11 isolates), while only 10% of RIF monoresistant and 5.61% of INH monoresistant showed additional FQ resistance.

On testing by Indirect DST which is the gold standard, FQ resistance among first line resistant isolates was also observed as 10%. Among the 11 resistant samples, three samples showed resistance to MFX (2.73%) at CB (indicating high level resistance), while, eight samples (7.27%) showed resistance to MFX at CC (indicating low level resistance).

### 3.3. Comparison of Direct and Indirect DST in terms of Time savings

Indirect DST required the initial culture step, total time taken to obtain the result was calculated by adding up the time taken for culture positivity since sample inoculation, time taken for setting up the indirect DST and time taken for obtaining the DST result. Hence, the turnaround time (TAT) for obtaining the results of indirect DST since the receipt of samples in the lab was on an average 25.6 ± 5 days (Table 1), with no correlation with respect to smear grading or resistance profile. In contrast, since direct DST do not require the initial culture step, time taken for it was calculated from the date of putting up of the DST till the reportable results were obtained. The average TAT for Direct DST was 15.59 ± 3.82 days with the time savings of 10 days ± 3.20 days over indirect DST (Table1). It also had statistically significant correlation with smear grading (Table 2) but insignificant correlation to resistance profile.

**Table 1.**
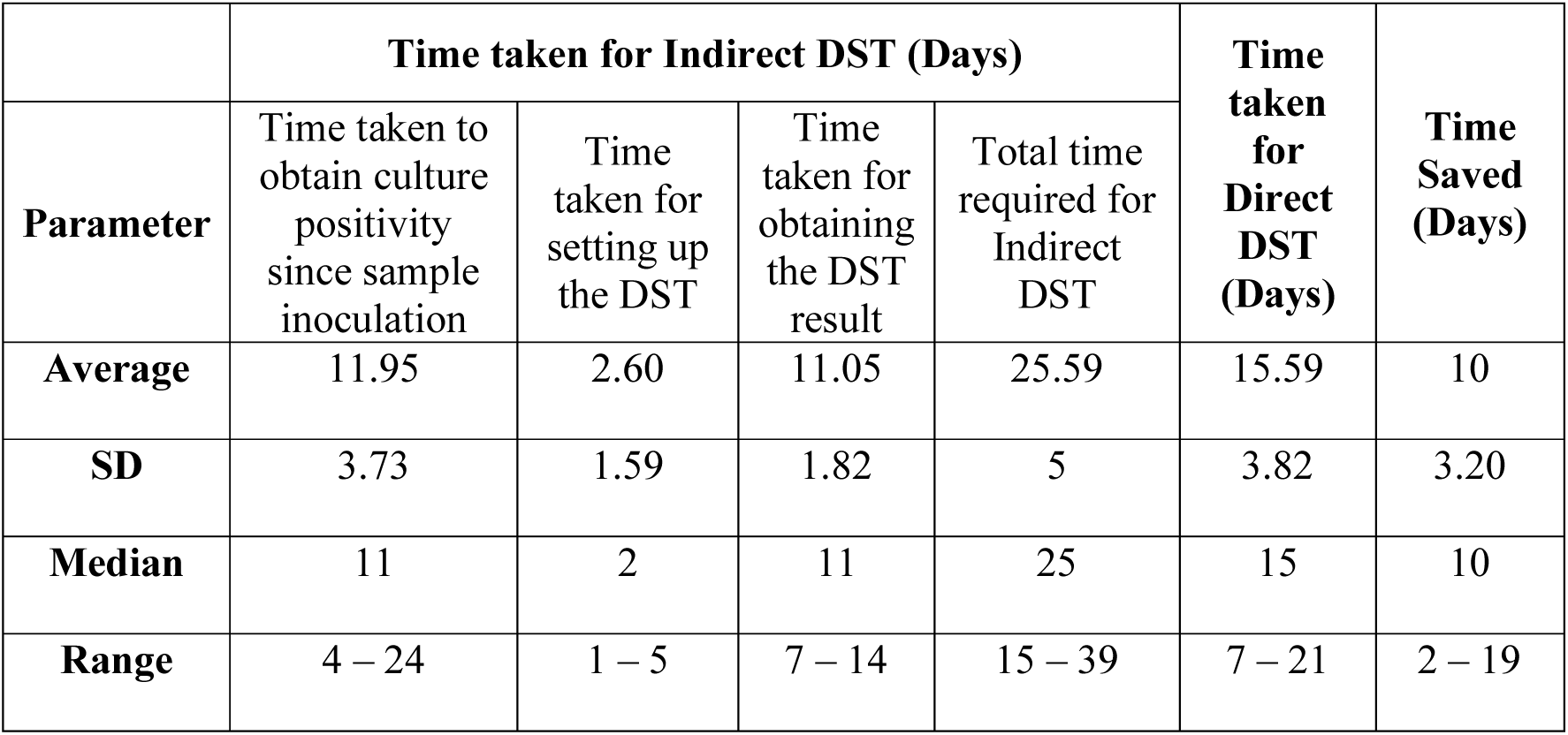
Time taken for Indirect and Direct DST method for MFX susceptibility testing.

**Table 2.**
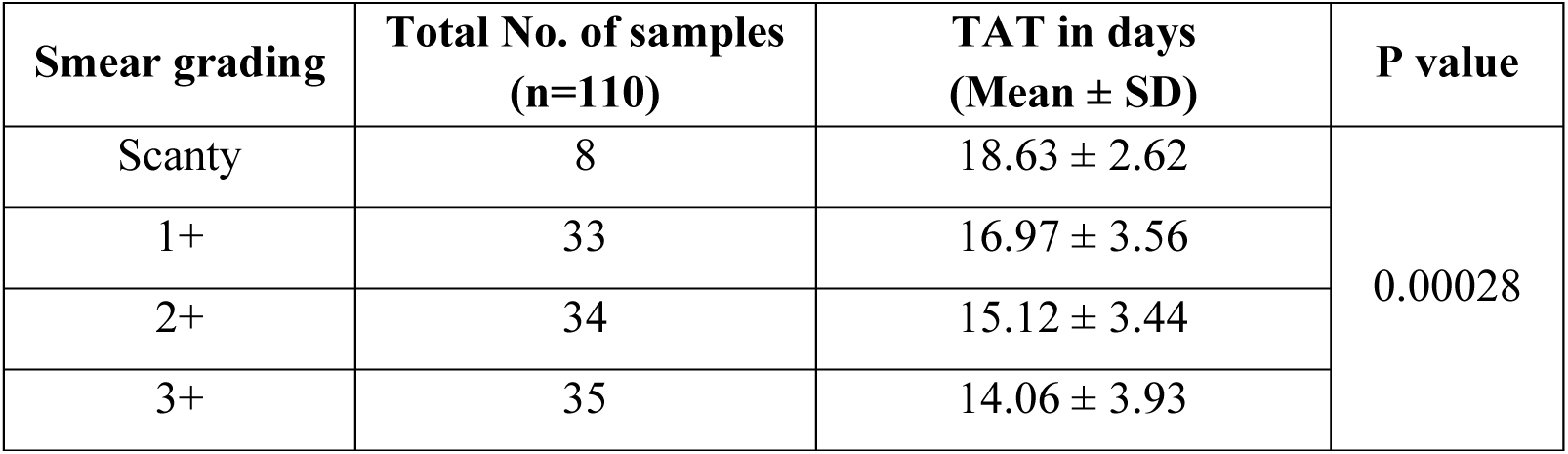
Time taken for Direct DST with respect to ZN smear grading.

### 3.4. Analysis for Diagnostic accuracy of Direct DST compared to Indirect DST and LPA

Direct DST reported MFX sensitivity in 90% (99 out of 110) and resistance in 10% (11 out of 110) samples. Among the resistant samples, based on the presence of growth at CC and/or CB, three samples (2.73%) had high level resistance while eight samples (7.27%) showed low level resistance to Moxifloxacin.

When compared with gold standard (Indirect DST), an excellent concordance of 98.18% (108 out of 110 samples) was noted between the methods, in terms of detection of drug resistance or susceptibility, giving a kappa value of 0.901 (Standard Error of kappa = 0.069; 95% confidence interval: From 0.765 to 1.000) indicating “Almost perfect agreement” between the methods. Discordance rate was obtained as 1.82% (2 out of 110) mainly with respect to identifying the strain as sensitive or resistant. (Table 3) However, there was no discrepancy in classifying the type of resistance (Table 4).

**Table 3.**
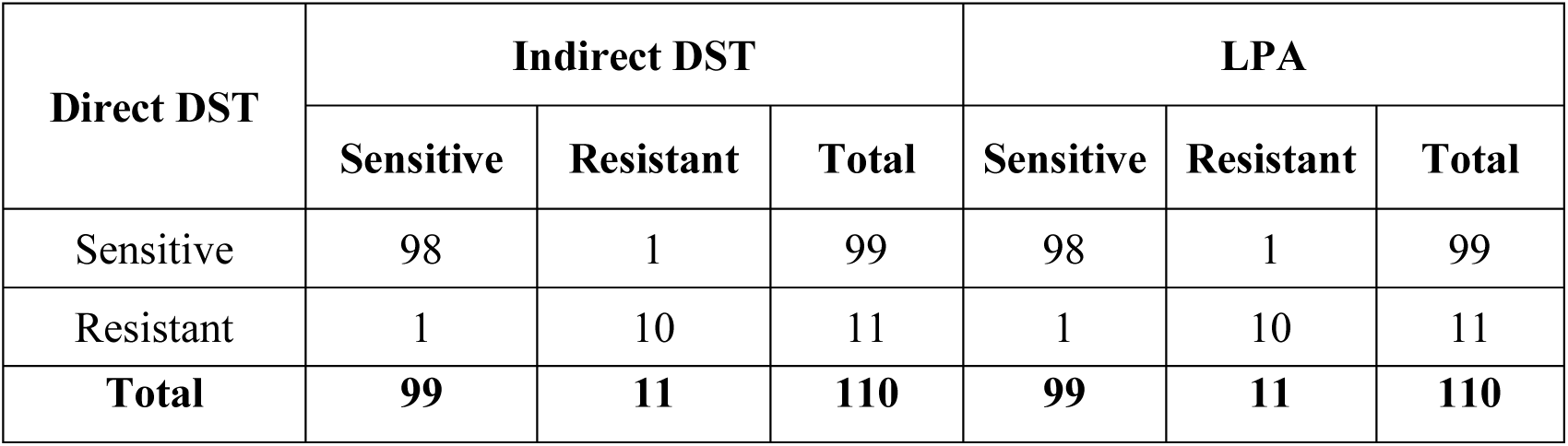
Comparison of MFX susceptibility results obtained by Direct with Indirect DST method and LPA.

**Table 4.** Ability of Direct DST to correctly classify High level and low level resistance with respect to Indirect DST and LPA.

|  |  | Indirect DST |  | LPA |  |
| --- | --- | --- | --- | --- | --- |
|  |  | High Level Resistance | Low Level Resistance | High Level Resistance | Low Level Resistance |
| **Direct DST** | High Level Resistance | 3 | 0 | 2 | 0 |
|  | Low Level Resistance | 0 | 7 | 3 | 4 |

Coincidentally, similar results were obtained when direct DST was compared with LPA with the exception of discrepancy of Direct DST in classifying the type of drug resistance identified by LPA. Direct DST was able to detect low level MFX resistance in all four cases which were showing resistance mutation conferring low level MFX resistance on LPA. However, it failed to identify high level resistance in three out of five cases (Table 4).

Similar discordance was also observed between Indirect DST and SL-LPA among 3 samples in classifying the type of resistance. Thus, diagnostic accuracy of Direct DST against the gold standard (Indirect DST) and LPA in identifying any kind of MFX resistance was similar as shown in table 5. However, in ability to correctly classify the level of resistance taking LPA as a reference, Direct DST showed a concordance of 95.45% (105 out of 110) resulting in “Substantial agreement” between the methods with a Cohen’s Kappa value of = 0.732 (Standard Error = 0.098; 95% Confidence interval: 0.540-0.923).

**Table 5.**
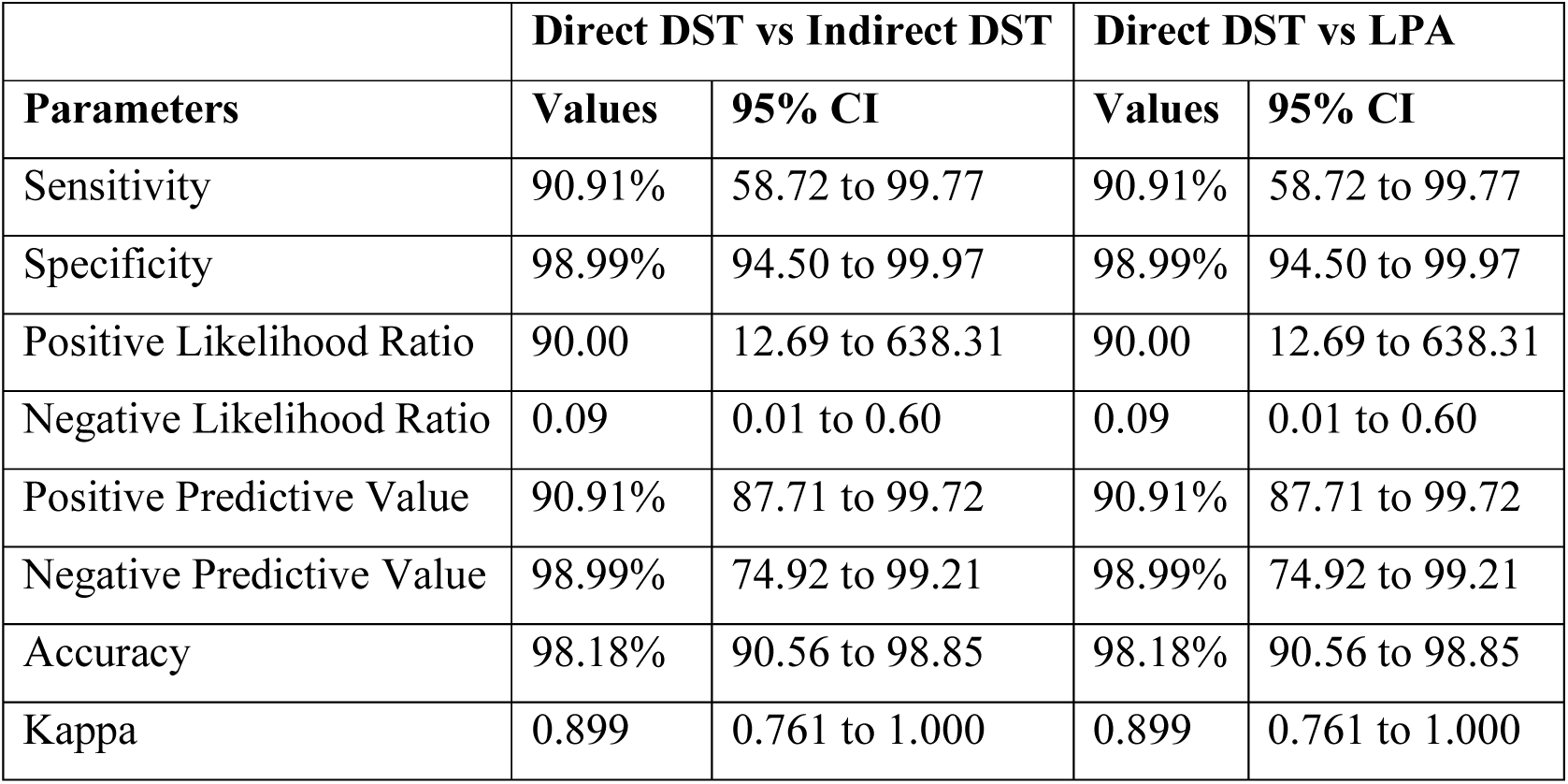
Diagnostic accuracy of Direct DST against Indirect DST and LPA.

### 3.5. Cost comparison of Direct and Indirect DST

The cost difference for processing of one sample by either of the methods was observed to be minimal (Approximately Rs. 341/-) and hence not considered as significant.

## 4. Discussion

Moxifloxacin is currently one of the most important drugs for treatment of DR-TB cases and has recently been introduced for treatment of DS-TB cases^5^. It has an efficacy comparable to isoniazid, hence cause rapid clearance of TB bacilli^2^. Considering the importance of the drug, it is placed under group A drugs for treatment of drug resistance TB^5^. According to the current diagnostic and treatment algorithm for TB followed in India, all RR/Hr/MDR-TB isolates are subjected to drug resistance testing by Second line-LPA to detect presence and type of resistance mutation, parallelly samples are subjected to Indirect DST using MGIT 960 system for moxifloxacin at CC and CB and other second line drugs. MFX phenotypic DST at two concentrations is essential to confirm the degree of resistance (low level or high level) indicated by SL-LPA. In case of low level MFX resistance, patient can be treated with MFX at higher doses (800 mg per day). Hence, treatment is started based on LPA results and modified after Phenotypic DST result is available. Indirect DST will take approximately 6-8 weeks to know the actual susceptibility pattern and thus delay the treatment decisions. Hence, our study tried to explore the performance of Direct DST method for moxifloxacin that involves DST directly from decontaminated sample without initial culture. Being a phenotypic DST, it can provide all the unique advantages of indirect DST but can also overcome the disadvantages of indirect DST by saving the crucial time.

Earliest protocols for direct DST exist for solid agar based media, though they have shown good performance for first line anti-TB drugs but such DST are much slower than MGIT 960 based DST^17,18^. Currently, only three other studies have tested the performance of direct DST on liquid culture- Siddiqi et. al study of 2012^13^, Zhang et. al. study of 2016^15^ and Srivastava et. al. study of 2025^19^.

A modified protocol compared to indirect DST was used in our study, where DST tubes were incubated for upto 21 days instead of 14 days in indirect DST, sample was diluted to 1:10 compared to 1:100 in growth control and ODAC-PANTA was used as supplement instead of ODAC supplement. This modified protocol was in line with Siddiqi et al. study which explored Direct DST for first line drugs and was taken as the reference for the current study^13^.

Similar protocol was also used by Srivastava et. al., however they followed the regular 14 day protocol instead of 21 days. Also, the tubes with X200 error obtained in their study were then observed for presence of flaky colonies which if present the tubes were then manually incubated for one more week and sensitivity was interpreted manually. This approach may result in human errors in interpretation of results^19^.

Our study tested Direct DST of Moxifloxacin at both CC (0.25 μg/ml) and CB (1.0 μg/ml) in contrast to Srivastava et. al. who tested Direct DST of moxifloxacin only at CC, though they additionally test levofloxacin at 1.0 μg/ml and other second line drugs.

Several studies concluded that moxifloxacin has unique advantages over levofloxacin- It has lower MIC, increased bactericidal activity, lower mutant prevention concentration and lower rate of resistance compared to levofloxacin^20–23^. It is also the only fluroquinolone that exhibit two level of resistance i.e. Low level and High level and hence a viable treatment option even for low level resistance at higher dose^9,12^. Studies also show that levofloxacin DST is a poor predictor of moxifloxacin resistance as both type of combinations- LVX-R+MXF-S and LVX-S+MXF-R were observed^24^.Hence DST of moxifloxacin at both CC and CB provides better picture of susceptibility to the drug.

In our study, time taken by indirect DST (average- 25.59 days) as well as Direct DST (Average - 15.59 days), was reported to be longer than other similar studies (Table 6). The higher time taken can be explained by the fact that, we received most samples from the other study site (Intermediate reference laboratory) after significant delay, hence samples had to be subjected to re-decontamination to overcome the contamination of the samples associated with prolonged storage, transportation and other routine lab work up on the patient sample. Despite this direct DST was associated with significant time savings compared to indirect DST and the time savings in our study was at par with other similar studies. An average of 10 days ± 3.2 days were saved in our case which was similar to Zhang et. al. study (Table 6). Moreover time taken by direct DST also reduced proportionately with the increased bacillary grading of the smear, this can further reduce the time required in cases with high bacterial burden.

**Table 6.** Time savings by Direct DST as reported by other published studies.

| **Study** | **Average time taken**<br>**for indirect DST** | **Average time taken**<br>**for direct DST** | **Time savings** |
| --- | --- | --- | --- |
| Present Study | 25.59 days | 15.59 days | 10 days |
| Siddiqi et. al.<sup>13</sup> | 19 days | 10-12 days | 6-9 days |
| Zhang et. al.<sup>15</sup> | 21.4 days | 10.9 days | 10.5 days |
| Srivastava et. al.<sup>19</sup> | 22 days | 10-12 days | 6-9 days |

When compared to Indirect DST which is the gold standard as well as LPA as a comparator, it reported similar sensitivity of 90.91%, specificity of 98.9% and overall accuracy of 98.18% with only one false sensitive and one false resistant result. It had an excellent concordance to indirect DST (98.18%) and “almost perfect agreement” with kappa value of 0.899. While, in terms of classifying the level of drug resistance, Direct DST performed slightly less accurate as compared to SL-LPA with Sensitivity, specificity of 90.91% and 98.99% respectively and a good concordance of 95.45% with “almost perfect agreement” with kappa value of 0.899 for detection of resistance. This is comparable to other similar studies for other antitubercular drugs^13,15,19^. Srivastava et. al. showed concordance-96.16%, sensitivity- 83.3%, specificity- 99.1% and accuracy- 98% for direct DST of moxifloxacin^13^, which was at par with our study.

Discordance though minimal were observed mainly in classification of resistance into low level and high level resistance between LPA and direct DST, three samples that were having high level resistance mutation by LPA, but exhibited low level resistance on Direct DST with similar discordance between LPA and indirect DST. This discrepancy between genotypic and phenotypic classification of resistance might be due to the strains having MICs higher than CC but lower than CB (between 0.25 to 1.0 μg/ml); other causes of phenotypic-genotypic discordance are- Bacilli with a resistance-conferring mutation present only in a minority of the bacterial population and are therefore picked up by sensitive molecular methods, but not in a sufficient percentage to be reliably detected by the proportion method^25^; mutations existing outside sequences targeted by genotypic tests like LPA that can also result in resistance^10,25,26^ ; Mutations that impart resistance also may reduce fitness of bacilli which may fail to grow on culture^27^; Genotypic assays cannot differentiate between living and dead bacilli and will show resistance mutation even in dead bacilli that will not grow in culture^28^.

Cost analysis showed no significant advantage of direct DST over indirect DST as resource utilisation is similar (MGIT tubes, supplement, falcon tube etc.), however using one 3-slot carrier instead of two PZA carrier will result in cost saving by reducing requirement of one MGIT tube and associated supplement for each sample. Advantage of Direct DST in terms of reduced labour requirement and reduced time requirement outweigh cost consideration. Primary drawbacks of the study were high error rates in terms of contamination (X400 error-7.63%) or failure of growth (X200 error- 8.39%), unequal representation by sensitive and resistant strains and minimal cost savings. We recommend further research to reduce the high error rates and to explore the possible reasons for Phenotypic – Genotypic discordance obtained by measuring the exact MICs of the strains leading to discordance and involvement of other molecular mechanisms that can be detected by sequencing.

The study concluded Direct DST to be a valid, accurate and time saving test for moxifloxacin susceptibility testing at both Critical concentration and Clinical Breakpoint when performed on smear positive sputum samples. It’s excellent performance characteristics for detection of resistance when compared to currently recommended methods like Indirect DST and SL-LPA makes it a reliable and rapid alternative to the gold standard saving significant time in guiding therapeutic decisions for effective patient management.

## 5. Conclusion

Direct moxifloxacin drug susceptibility testing against Mycobacterium tuberculosis performed on sputum samples demonstrated excellent performance characteristics in terms of accuracy and time savings as compared to the gold standard. Therefore, it can prove to be a reliable and rapid alternative to the current approach of waiting for the results of indirect phenotypic moxifloxacin drug susceptibility testing to ensure the therapeutic efficacy of moxifloxacin at clinical breakpoints in patients showing low-level resistance mutations on genotypic testing. This in turn will save a significant amount of time and resources in guiding therapeutic decisions for efficient patient management within programmatic frameworks.

## 6. Abbreviations

CB: Clinical Breakpoint
CC: Critical Concentration
DR-TB: Drug Resistant TB
DST: Drug susceptibility testing
FL-LPA: First Line-Line Probe Assay
FQ: Fluroquinolone
Hr-TB: Isoniazid-resistant TB
INH: Isoniazid
LC-DST: Liquid Culture Drug susceptibility testing
LPA: Line Probe Assay
MDR-TB: Multidrug-resistant tuberculosis
MFX: Moxifloxacin
MGIT: Mycobacterium growth indicator tube
MTB: Mycobacterium tuberculosis
MTBC: Mycobacterium tuberculosis complex
NTEP: National Tuberculosis Elimination Programme
RIF: Rifampicin
SL-LPA: Second Line-Line Probe Assay
WHO: World Health Organization

## 7. Acknowledgement

We thank the National TB Elimination Program (NTEP), Government of India for their valuable support in providing the drug powders as well as clinical material for this study.

## 8. Author Contributions

Subhasish Bhadra – Conceptualization, Data Curation, Formal analysis, Investigation, Methodology, Visualization, Writing – original draft, Writing – Review & Editing; Ujjwala Gaikwad – Conceptualization, Formal analysis, Investigation, Methodology, Visualization, Project Administration, Resources, Supervision, Validation, Writing – original draft, Writing – Review & Editing; Sachin Chandrakar – Investigation, Methodology, Project Administration, Resources; Kumar Vikram and Abhijit Prasad - Supervision, Writing – review and editing.

## 10. Data availability

The datasets used and/or analysed during the current study are available from the corresponding author on reasonable request.

## 11. Declarations

### 11.1. Ethical approval and consent to participate

The study was conducted only after obtaining written informed consent from the patients for their participation in the study. A written informed consent to participate was obtained from the Parents or Legal guardians of the participants under the age of 18 years, additionally a written assent was obtained from the participants belonging to the age group of 12-18 years.

Consent waiver was approved by the Institute Ethics Committee of the All India Institute of Medical Sciences, Raipur (Chhattisgarh) for the samples received from IRL, Lalpur vide. 4832/IEC-AIIMSRPR/2024.

This study has been ethically approved by the Institute Ethics Committee (4088/IEC-AIIMSRPR/2023) of the All India Institute of Medical Sciences, Raipur (Chhattisgarh).

### 11.2. Consent for publication

Not applicable.

### 11.4. Clinical Trial Number

Not Applicable

